# Early Emergence of Abnormal Muscle Synergies in the Human Upper Extremity Following Stroke

**DOI:** 10.64898/2026.08.19.26360812

**Authors:** Abed Khorasani, Cynthia M. Gorski, Vivek Paul, Na-Teng Hung, Joel Hulsizer, Prashanth R. Prakash, Fan Caprio, Richard L. Harvey, Jinsook Roh, Marc W. Slutzky

## Abstract

**Background:** Abnormal muscle co-activation, also called abnormal synergies, is an important contributor to arm impairment after stroke. While abnormal co-activation is well-described in chronic stroke, it remains unclear how early abnormal patterns appear and whether their spatial and temporal characteristics resemble those seen in the chronic phase. We sought to determine how soon after stroke abnormal muscle co-activation appears.

**Methods:** In this cross-sectional study, thirty-nine individuals with hemiparesis in the early subacute period (<21 days) and sixty-eight individuals in the chronic period (>6 months) after stroke performed targeted reaching movements while surface electromyography (EMG) was recorded from nine upper-limb muscles. Muscle synergies (patterns of coordinated muscle activation) were identified using non-negative matrix factorization. Synergy composition (spatial structure) and activation profile (temporal structure) were compared across the contralesional arms of subacute and chronic participants and the ipsilesional arm, which served as the reference for normal coordination.

**Results:** Three primary synergies accounted for most EMG variance during reaching in each arm group. A deltoid-dominant synergy characterized by abnormal co-activation of anterior and posterior deltoids, was present in both subacute and chronic stages in the contralesional arm but was absent in the ipsilesional arm. In addition, the elbow flexor synergy co-activated with the deltoid synergy in both contralesional groups but not in the ipsilesional arm. Abnormal co-activation between elbow flexor and elbow extensor synergies was also seen in contralesional, but not ipsilesional, arms. These abnormalities were already present by 15 days after stroke and did not differ between subacute and chronic groups.

**Conclusions:** Abnormal muscle co-activation appears within the first few weeks after stroke and persists in chronically impaired survivors. Its full development this early suggests these patterns arise rapidly rather than emerging gradually during recovery, and that interventions targeting abnormal co-activation may be most useful when applied early.

**Clinical Trial Registration:** NCT03401762.

## Introduction

Stroke disrupts the neural circuits that coordinate multi-joint arm movements and leads to impairments such as weakness, loss of dexterity, spasticity, and abnormal patterns of muscle co-activation known as abnormal synergies [1], [2], [3]. These abnormal synergies limit the ability to generate flexible and independent joint movement and are correlated with the severity of upper-limb motor impairment after stroke [4], [5], [6].

Abnormal joint coupling has long been described in the chronic phase after stroke [1], [3]. In stroke survivors with severe arm impairment, abnormal co-activation was prominent among proximal muscles, including excessive coupling between the anterior and posterior deltoids during both free and isometric reaching [6], [7]. Despite this detailed characterization in chronic stroke, the onset and progression of these abnormal coordination patterns during the initial weeks after stroke are still not well quantified.

The term “synergy” has been used clinically to describe both the abnormal joint coupling after stroke [3] and the coordinated patterns of muscle activity, whether in neurotypical or impaired limbs. For clarity, here we use synergy to mean coordinated pattern of muscle activity and specify whether these patterns are abnormal or normal. Abnormal co-activation after stroke can manifest in two ways. First, the wrong muscles may be grouped together within a synergy — for example, anterior and posterior deltoids being in a single module when they should activate independently [6], [8] (abnormal synergy composition). Second, synergies that should activate at different times in a movement may instead activate at the same time (abnormal co-activation between synergies).

Classical observations by Twitchell described the appearance of abnormal flexor (shoulder, elbow, and wrist flexion) and extensor (shoulder and elbow extension) co-activation patterns within the first two months after stroke onset [3]. However, this study was performed in a population with unclear but seemingly broad range of arm impairment outcomes, did not quantify the patterns, and listed a wide range of onset times of these patterns. A recent quantitative kinematic study showed that even within 8 weeks of stroke, survivors already exhibit altered shoulder–elbow coordination and intrusion of flexor-dominated patterns during reaching movements [9]. Within two weeks of a cortical lesion in rodents, the forelimb displayed disrupted proximal–distal coordination, reduced movement smoothness, and altered joint angles [10]. Together, these findings suggest that abnormal co-activation may develop very early; however, the precise timing, the specific types of co-activation patterns, and how these early patterns compare to those observed in the chronic phase remain unclear.

Here, we aimed to address this gap by examining both the composition and activation timing of muscle synergies during reaching in the contralesional arms of subacute and chronic stroke survivors. We compared these to those in the ipsilesional arm to identify deviations from typical coordination patterns. We found evidence that abnormal co-activation patterns observed in the first few weeks resemble those present years later. Understanding the early emergence of these patterns is critical for guiding the timing of interventions aimed at preventing or attenuating abnormal co-activation and improving motor recovery.

## Methods

### Participants

Participants in the subacute group were screened within 21 days of stroke onset. Eligibility criteria included being at least 18 years old, having a first-ever unilateral ischemic or hemorrhagic stroke, and a SAFE (Shoulder Abduction–Finger Extension) score of 1-8 at the first evaluation [11]. Exclusion criteria for the subacute group were the following: prior stroke, other neurological disorders affecting the upper limb, severe cognitive or language impairments preventing understanding of task instructions, major sensory deficits affecting the paretic limb, severe pain or orthopedic conditions limiting upper-limb movement. Muscle synergy analysis (see below) was performed only when at least 6 of the 9 recorded upper-limb muscles showed activation during the task, ensuring sufficient numbers of EMGs to perform reliable muscle synergy extraction.

The current study involved a subset of participants enrolled in a randomized clinical trial of wearable myoelectric interface for neurorehabilitation for arm function recovery [12]. Participants in the chronic group were included if they had a first-time, unilateral stroke at least six months prior to screening and had moderate-to-severe upper extremity impairment, defined by a Fugl-Meyer Assessment of the Upper Extremity (FMA-UE) score of less than 30 [13]. Exclusion criteria for the chronic group were the following: severe visual impairment, significant language comprehension deficits preventing understanding of task instructions, prior stroke, fixed contractures in the affected upper limb, botulinum toxin injections in the affected arm within the preceding three months, starting a new physiotherapy program in the past three months, or participation in another research study involving the upper limb in the past three months.

The ipsilesional arm of a subset of participants from both the subacute and chronic cohorts served as a control arm. This subset was selected primarily due to time constraints and the retrospective availability of usable data from chronic participants.

The subacute contralesional arm, chronic contralesional arm, and ipsilesional arm groups were matched on age and impairment level based on each participant’s FMA-UE score at the time of synergy analysis, ensuring comparable functional status across cohorts.

This study was conducted at Northwestern University and the Shirley Ryan AbilityLab in Chicago. All procedures were approved by the Northwestern University Institutional Review Board (NCT03401762), and all participants provided written informed consent. This study is reported in accordance with the STROBE guidelines (Strengthening the Reporting of Observational Studies in Epidemiology).

### Clinical assessment and EMG Recording

Trained occupational therapists measured arm functional activity with the Wolf Motor Function Test (WMFT), motor impairment with FMA-UE, and spasticity evaluated at the wrist, elbow, and shoulder joints with the Modified Ashworth Scale (MAS) in all participants. Occupational therapists were trained by experienced therapists to ensure consistent administration of assessments, achieving over 95% agreement in FMA-UE performed on a sample participant with stroke, graded by the training therapist.

Participants were seated with their body stabilized and executed reaches to three different targets located in front of (sagittal plane) and to the side of (coronal plane) the arm at both shoulder and hip height (Fig. 1A). We recorded surface EMG from nine arm muscles: brachioradialis (BRD), biceps brachii (BI), triceps brachii long head (TRIlong), triceps brachii lateral head (TRIlat), anterior deltoid (AD), middle deltoid (MD), posterior deltoid (PD), pectoralis major (Pec), and trapezius (Trap). Placement followed SENIAM standards, with amplification via the Delsys Trigno system [14].

**Figure 1.**
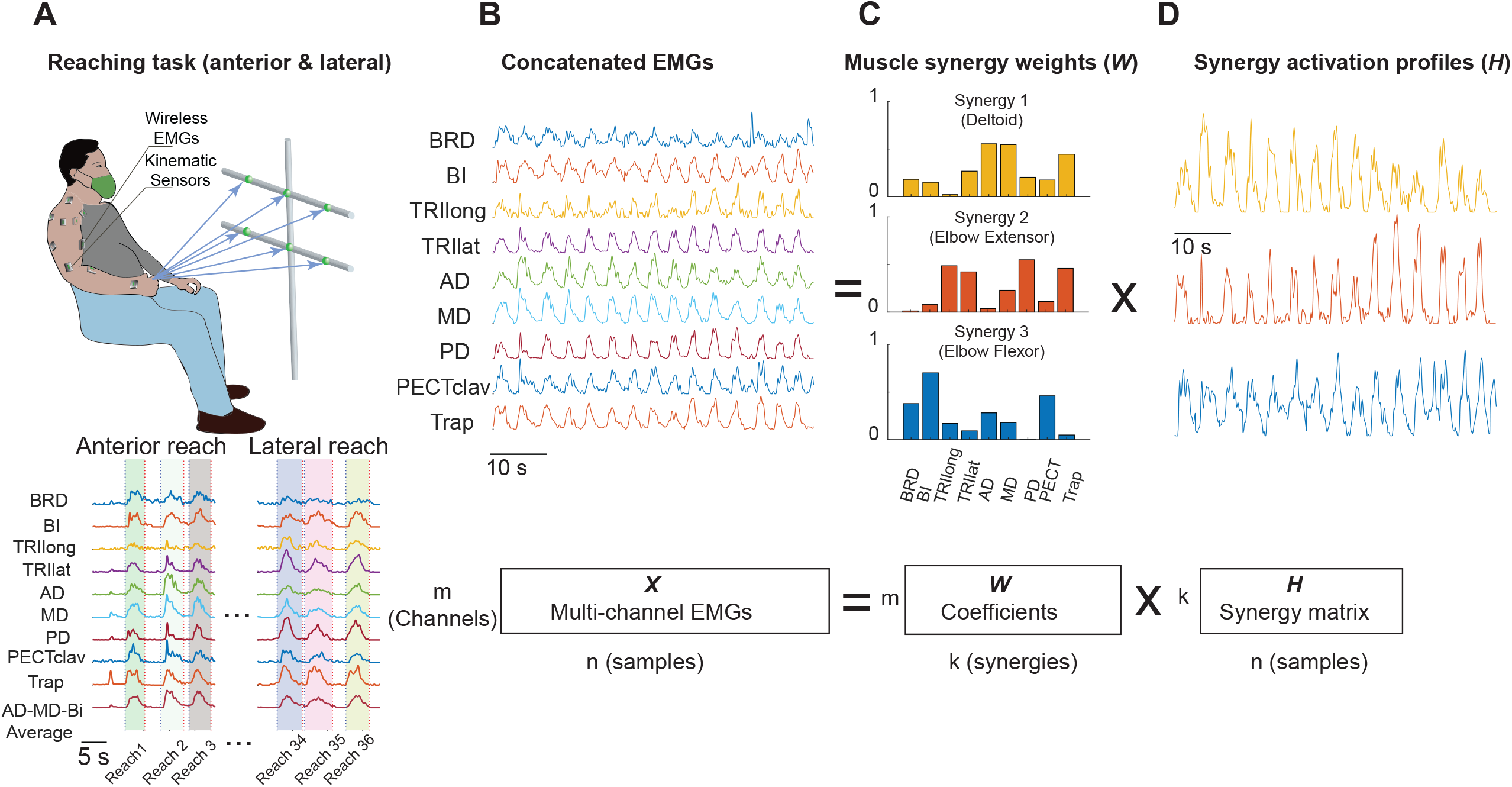
Reaching task, EMG recordings, and muscle synergy identification. **A**, Participants were seated and performed repeated anterior and lateral reaching movements to targets mounted on a vertical frame set at shoulder and waist heights. Surface EMGs were recorded from nine upper-limb muscles. The targets required participants to initiate the reach from a neutral starting position and extend the arm in first the sagittal, then coronal, plane. **B**, Representative concatenated EMG envelopes from a full sequence of reaches. EMG signals were recorded from brachioradialis (BRD), biceps brachii (BI), triceps long head (TRIlong), triceps lateral head (TRIlat), anterior deltoid (AD), middle deltoid (MD), posterior deltoid (PD), clavicular pectoralis (PECclav), and trapezius (TRP). **C**, Muscle synergy weight vectors (W matrix) obtained from non-negative matrix factorization of the concatenated EMG envelopes. Three synergies accounted for more than 90% of the variance: the deltoid synergy (Synergy 1, yellow), the elbow extensor synergy (Synergy 2, orange), and the elbow flexor synergy (Synergy 3, blue). Each bar represents the relative weight of an individual muscle within a given synergy vector. The mathematical relationship between the multi-channel EMG matrix, the synergy weight matrix W, and the activation profile matrix H is illustrated in the schematic below. **D**, Synergy activation profiles (H matrix) across the concatenated reaching sequence for the three synergies (colors as in C). Each trace represents the time-varying activation coefficient of a given synergy across successive reaches, demonstrating structured temporal modulation of each synergy module during movement.

### Data processing and synergy analysis

Surface EMGs were recorded at 2 kHz, band-pass filtered (50–450 Hz, 4th-order Butterworth, zero-phase), full-wave rectified, and then low-pass filtered at 5 Hz with the same filter type and order to obtain EMG envelopes. Envelopes were downsampled to 20 Hz and segmented from 0.5 s before movement onset to 0.5 s after movement offset (Fig. 1A). Movement onset and offset were determined from the average activation of AD, BI, and MD muscles, which consistently exhibited robust activity across participants.

Segmented trials were concatenated to form an EMG data matrix (m × n; where m was the number of muscles and n was the total number of samples). Each channel was normalized to its maximum value across all trials within each session to minimize bias from amplitude differences. Muscle synergies were identified using non-negative matrix factorization, which decomposes the EMG matrix X(t) (Fig. 1B) into two non-negative matrices:

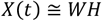

where *W* (m × k) contains time-invariant synergy vectors (Fig. 1C) and *H* (k × n) contains their time-varying activation profiles (Fig. 1D). The number of synergies (k) was determined by variance accounted for (VAF) criteria: global VAF (in EMGs from all muscles) > 90% and an increase in VAF of less than 5% with the addition of an extra synergy [6], [7], [15]. Based on these criteria, three synergies were retained across groups to facilitate comparisons among the three arm groups: ipsilesional, subacute contralesional, and chronic contralesional. These three synergies were consistently identified as deltoid-dominant synergy (Delt), an elbow extensor synergy (EE), and an elbow flexor synergy (EF).

To ensure consistent synergy labeling across participants, group-average synergy vectors were first computed separately for each arm group by averaging synergy vectors across participants in that group. For each participant arm, the extracted synergy vectors were then matched to the group-average vectors for each of the 3 synergies by selecting the group-average vectors with the highest Pearson correlation coefficient (R) with each individual synergy vector.

To determine an objective similarity criterion, a permutation analysis was performed. Specifically, 1,000 surrogate synergy sets were generated by randomly shuffling the muscle weights within each extracted synergy matrix. Pairwise Pearson correlation coefficients were computed between all surrogate synergy pairs, and the 95th percentile of the resulting null distribution (R = 0.75) was used as the minimum threshold for accepting a synergy as a valid match to the norm template. Participants whose individual synergy fell below this threshold for a given synergy were excluded from analyses involving that synergy.

### Synergy composition similarity

To compare synergy composition across groups, the mean (norm) synergy weight vectors for each synergy (Delt, EE, EF) were computed across all ipsilesional arms and served as reference templates. For each participant arm in all three groups (ipsilesional, subacute contralesional, and chronic contralesional), the Pearson correlation coefficient (R) between each participant arm’s individual synergy weight vector and the corresponding ipsilesional norm vector was computed separately for each synergy. This yielded one R value per participant arm per synergy, reflecting how closely that participant arm’s synergy composition matched the ipsilesional reference. R values were compared across groups using one-way ANOVA with Tukey–Kramer post-hoc tests (α = 0.05).

### Synergy temporal coupling

To quantify the relative timing of synergy activations during reaching, the Pearson correlation coefficient was computed between the mean activation profiles of each of three synergy pairs within each participant: EF and EE, EF and Delt, EE and Delt. To account for differences in movement duration, we normalized the timing by linearly interpolating each trial to a fixed length of 101 samples, representing 0–100% of the movement duration. The time-normalized activation profiles were then averaged within each participant before correlating. A correlation near zero indicated that two synergies activated independently and at distinct phases of the reach.

### Statistical Analysis

All eligible participants with usable data over the enrolment period were included; no a priori power calculation was performed. Statistical analyses were performed separately for synergy composition and synergy temporal coupling.

Group differences in synergy composition similarity and synergy temporal coupling were assessed using one-way ANOVAs with Tukey–Kramer post-hoc tests (α = 0.05). For synergy composition, R values representing the similarity between each participant arm’s synergy vector and the ipsilesional norm template were compared for each synergy across the three arm groups. For temporal coupling, pairwise correlation coefficients between synergy activation profiles were compared across groups for each pair (EF vs. EE, EF vs. Delt, EE vs. Delt).

## Results

In this study, 39 participants with subacute stroke were included, ranging in age from 26 to 91 years, with a mean FMA-UE score of 21.0 ± 10.6 (mean ± SD). The subacute group was evaluated 15 ± 8 (mean ± SD) days after stroke onset. The chronic cohort consisted of 68 participants aged 20–87 years, with an average FMA-UE score of 19.5 ± 8.1 and a mean time post-stroke of 1934 ± 2000 days (5.9 ± 7.3 years). EMGs were recorded in the ipsilesional arm in 12 participants (4 subacute and 8 chronic). Demographic and clinical characteristics are presented in Table 1. There were no significant differences in age (p = 0.12) or FMA-UE (p = 0.18) among groups.

**Table 1.** Participant Demographics.

|  | **Chronic Stroke**<br>**(n=68)** | **Subacute Stroke**<br>**(n=39)** | **p-value** |
| --- | --- | --- | --- |
| **Age (years)** | 57.8 ± 14.1 | 62.3 ± 14.4 | 0.12 |
| **Gender, n (%)** |  |  |  |
| **Male** | 45 (66) | 18 (54) |  |
| **Female** | 23 (34) | 21 (46) |  |
| **Race, n (%)** |  |  |  |
| **White** | 35 (51) | 17 (44) |  |
| **Black** | 26 (38) | 16 (41) |  |
| **Asian** | 5 (8) | 3 (8) |  |
| **Other** | 2 (3) | 3 (8) |  |
| **Affected side** |  |  |  |
| **Right** | 35 (51) | 10 (26) |  |
| **Left** | 33 (49) | 29 (74) |  |
| **Time since stroke** | 5.9 ± 7.3 (yrs) | 15.2 ± 8.1 (days) | <0.001 |
| **FMA-UE (out of 66)** | 19.5 ± 8.1 | 21.0 ± 10.6 | 0.18 |
| **WMFT (s) (out of 120)** | 82.2 ± 25.2 | 85.9 ± 28.5 | 0.51 |
| **MAS (out of 4)** | 0.8 ± 0.4 | 0.2 ± 0.2 | <0.001 |

Clinical (FMA-UE, WMFT, and MAS) and demographic values (age, time since stroke) are presented as mean ± SD. WMFT: Wolf Motor Function Test, FMA-UE: Fugl-Meyer Assessment of the upper extremity, MAS: Modified Ashworth Scale.

In all three groups, NMF consistently identified three dominant synergies that together captured most (90%) of the total EMG variance. For the subacute contralesional arms, chronic contralesional arms, and ipsilesional arms, the number of synergies required to explain at least 90% of the variance was 3.4 ± 1.0, 2.7 ± 0.9, and 3.4 ± 0.5 (mean ± SD), respectively. To enable direct comparison, we retained three synergies for all arm groups, resulting in global VAF values of 90.9 ± 3.6, 92.5 ± 2.7, and 90.9 ± 1.9, respectively.

### Synergy composition is altered early after stroke

Analysis of synergy composition (synergy weight patterns) revealed marked abnormalities in both subacute and chronic contralesional arms compared to the ipsilesional arm (Fig. 2A). The most notable finding was the presence of strong co-activation of anterior and posterior deltoids (AD-PD) and the trapezius (TrP) in the Deltoid synergy. This key deviation from the normal pattern was present in both subacute and chronic contralesional arms but not in ipsilesional arms and was detectable within the first 2-3 weeks after stroke. In addition to the Deltoid synergy, composition of the other two synergies was altered early after stroke. In Synergy 2, the elbow extensor synergy, contralesional arms had lower weights for middle and posterior deltoids (MD-PD) than in the ipsilesional arm. Synergy 3, the elbow flexor synergy, showed decreased trapezius (Trp) weight in subacute and chronic contralesional arm. Ipsilesional participants showed consistently high R values across all three synergies (deltoid: R = 0.91 ± 0.03; EE: R = 0.94 ± 0.04; EF: R = 0.93 ± 0.05), confirming that the ipsilesional norm template reliably captures typical synergy composition.

**Figure 2.**
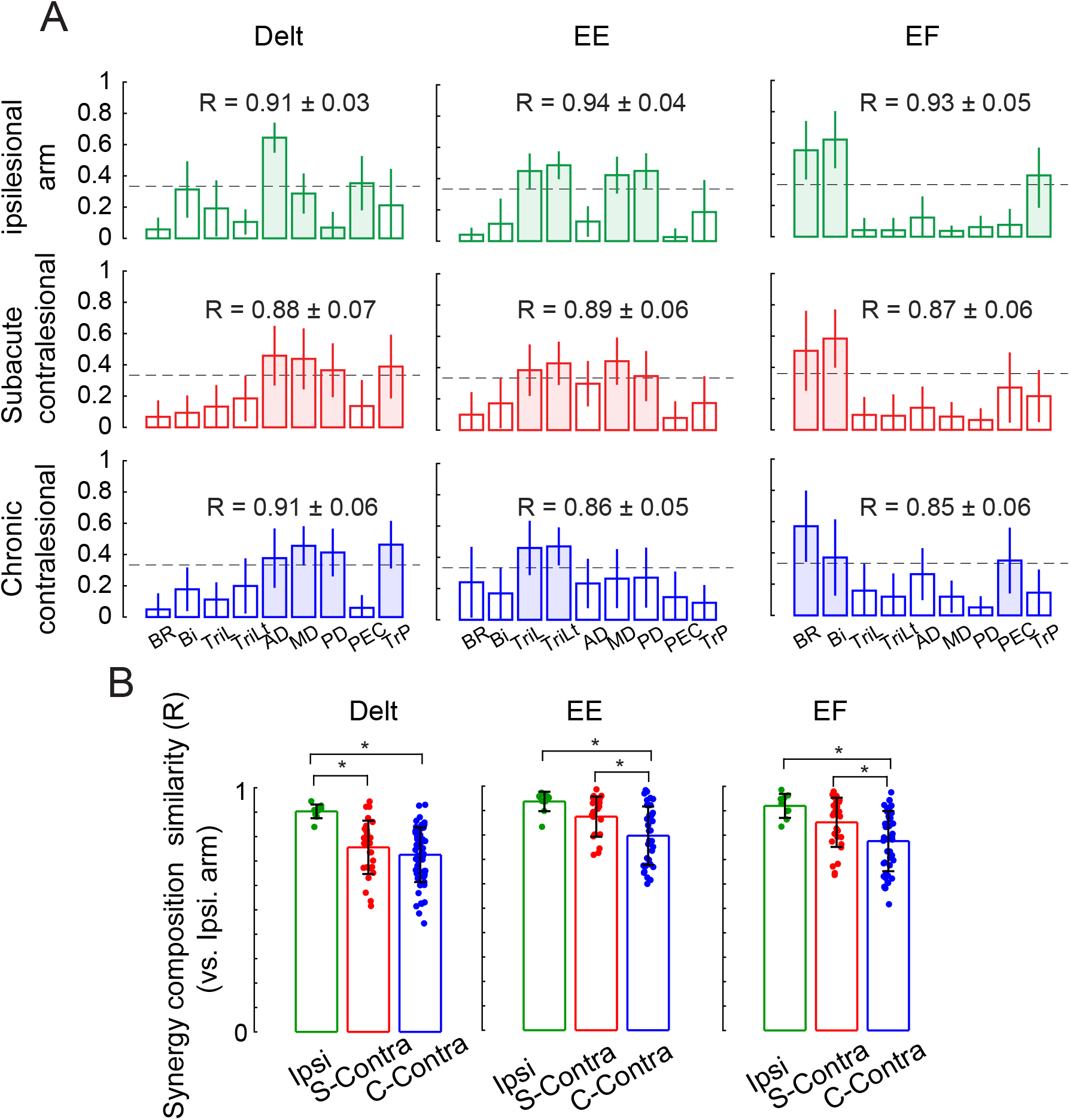
Muscle synergy compositions and similarity across contralesional and ipsilesional arms. **A**, Mean (± SD) muscle synergy vectors for the deltoid (Delt), elbow extensor (EE), and elbow flexor (EF) synergies identified in each arm group: ipsilesional arm (top, green), subacute contralesional arm (middle, red), and chronic contralesional arm (bottom, blue). Each bar represents the weight of a given muscle within a synergy vector. Filled (shaded) bars indicate muscles whose weights exceeded the co-activation threshold (dashed horizontal line). Muscle abbreviations as in Fig. 1. **B**, Synergy composition similarity (R) of each participant’s synergy vector relative to the ipsilesional norm template — the mean synergy vector computed across all ipsilesional arms — shown separately for the Delt, EE, and EF synergies. Each dot represents one participant arm’s R value. Bar height and error bars indicate group mean ± SD. R values were compared across all three groups using a one-way ANOVA with Tukey-Kramer post-hoc tests. Asterisks indicate significant pairwise differences (*, p < 0.05). Ipsi: ipsilesional arm; S-Contra: subacute contralesional arm; C-Contra: chronic contralesional arm.

Correlations with the ipsilesional norm differed significantly across groups for all three synergies (one-way ANOVA, all p < 0.001; Fig. 2B). For the deltoid synergy, both the subacute and chronic contralesional arms had significantly lower R values than the ipsilesional group (p = 0.004 and p = 5×10^−6^). For the EE and EF synergies, only the chronic contralesional arm showed significantly lower R values than the ipsilesional group (p = 0.0004 and p = 0.002), while the subacute contralesional arm did not differ from the ipsilesional group (p = 0.2 and p = 0.3). The two contralesional groups differed significantly from each other for both synergies (p = 0.01 and p = 0.02).

### Elbow flexor synergy co-activates abnormally with deltoid and elbow extensor synergies in the contralesional arm starting in the subacute stage

The temporal structure of synergy activations during reaching differed markedly between the ipsilesional and contralesional arms (Fig. 3A). In the ipsilesional arm, the deltoid synergy activated early in the reach, whereas the elbow flexor synergy activated later, resulting in clearly separated temporal profiles (Fig. 3A, left). In contrast, in both the subacute and chronic contralesional arms, the elbow flexor and deltoid synergies activated early in the reach, at nearly the same time, producing overlapping profiles throughout the reach (Fig. 3A, middle and right). This suggests that the normal independent activation of shoulder and elbow muscle synergies is disrupted very early after stroke.

**Figure 3.**
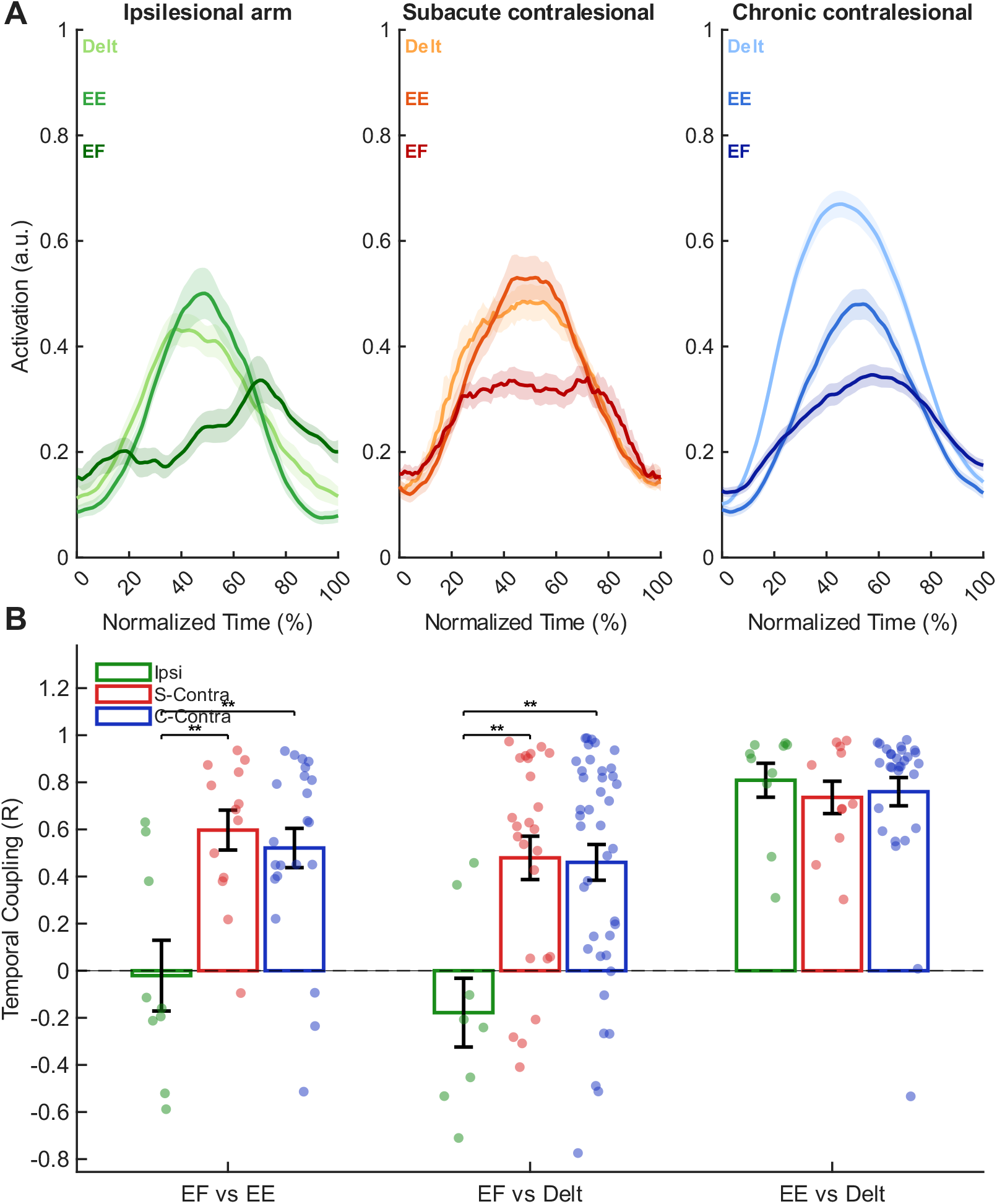
Elbow flexor synergy co-activated abnormally with the other synergies in the contralesional arm groups. **A**, Group-averaged, time-normalized activation profiles for the deltoid (Delt), elbow extensor (EE), and elbow flexor (EF) synergies during reaching, shown separately for the ipsilesional arm (left, green), subacute contralesional arm (middle, red/orange), and chronic contralesional arm (right, blue). Within each group, each synergy activation profile is represented by a distinct shade: Delt (lightest), EE (medium), and EF (darkest). In the ipsilesional arm, synergy modules activate in temporally distinct phases, with Delt peaking early and EF activating later in the reach. In both contralesional groups, EF and Delt show increased temporal overlap throughout the reach. Shaded regions represent ± standard error of the mean. **B**, Pairwise Pearson correlation coefficients (R) between synergy activation profiles, computed within each participant (dots) and compared across groups. Three synergy pairs are shown: EF vs EE, EF vs Delt, and EE vs Delt. Each bar represents the group mean ± SEM. Brackets indicate significant between-group differences (**, p < 0.01; Tukey-Kramer post-hoc test). Only participants whose synergy composition similarity exceeded the threshold (R ≥ 0.75) for both synergies in a given pair were included in that pair’s analysis. Ipsi = ipsilesional arm; S-Contra = subacute contralesional arm; C-Contra = chronic contralesional arm.

In the ipsilesional arm, the elbow flexor and elbow extensor synergies were temporally uncorrelated (R = −0.02 ± 0.1), reflecting their distinct activation timing during reaching (Fig. 3B, left). Both the subacute and chronic contralesional arms showed significantly higher EF–EE correlations (R = 0.6 ± 0.08 and R = 0.5 ± 0.08, respectively), indicating that these two synergy activations became abnormally coupled after stroke. These values were significantly higher than that in the ipsilesional arm (Ipsi vs. S-Contra: p = 0.002; Ipsi vs. C-Contra: p = 0.002), while the subacute and chronic groups’ correlations did not differ from each other (p = 0.8).

A similar pattern was observed for the elbow flexor and deltoid synergy pair. The ipsilesional arm showed no significant temporal correlation (R = −0.2 ± 0.1; Fig. 3B, middle). Both contralesional groups showed significantly higher correlations (S-Contra: R = 0.5 ± 0.09; C-Contra: R = 0.5 ± 0.08) than the ipsilesional arm (Ipsi vs. S-Contra: p = 0.003; Ipsi vs. C-Contra: p = 0.002), while the subacute and chronic groups did not differ significantly from each other (p = 1). The elbow extensor and deltoid synergy activation patterns were strongly correlated in all three groups (Ipsi: R = 0.8 ± 0.07; S-Contra: R = 0.7 ± 0.07; C-Contra: R = 0.8 ± 0.06; Fig. 3B, right). No significant between-group differences were observed (all p > 0.8).

Taken together, these findings show that stroke specifically disrupts the temporal independence of the elbow flexor synergy, which becomes abnormally coupled with both elbow extensor and deltoid synergies. This abnormal coupling was already fully present in the subacute stage.

## Discussion

This study sought to determine how early after stroke abnormal co-activation appears and whether abnormal muscle synergy patterns in the subacute phase resemble those seen in the chronic phase. We found two abnormal patterns in muscle synergy composition and temporal activation were already fully developed by a mean of 15 days after stroke. First, the deltoid-dominant synergy included abnormal co-activation of anterior and posterior deltoids, as well as trapezius. Second, the elbow flexor synergy co-activated with the deltoid and elbow extensor synergies during reaching. Together, these findings suggest that abnormal co-activation, both composition and timing, is not a late compensation but rather an early contributor to impaired arm control after stroke. Together, these two forms of abnormal co-activation reproduce, at the level of muscle activity, the abnormal shoulder–elbow coupling that Twitchell [3] described clinically and Dewald and colleagues quantified mechanically [4].

### Abnormal synergy composition and temporal coupling are present early after stroke

The abnormal deltoid and trapezius co-activation pattern mirrors our previous work in severely impaired chronic stroke survivors, in which excessive coupling of anterior and posterior deltoids was seen during both free and isometric reaching, and was strongly associated with impaired kinematics [6], [7], [15]. Importantly, the present data extend those prior observations by demonstrating that impaired deltoid fractionation is present almost as soon as voluntary EMG reappears after stroke, implying that the window for preventing abnormal co-activation may be very narrow and occur very early in recovery.

Abnormal temporal coupling between synergies represents a phenomenon that is distinct from changes in synergy composition. Abnormal composition reflects a reduced ability to independently activate muscles that become incorporated into a common module. In contrast, abnormal temporal coupling occurs when individual synergies remain identifiable but are recruited simultaneously. Existing reports of post-stroke synergy abnormalities, including synergy merging and impaired fractionation [16], primarily address changes in synergy composition and do not fully explain this temporal loss of independence.

### Relationship to prior studies of early recovery

Abnormal synergy composition and temporal coupling were already present at approximately two weeks after stroke. This might suggest that these abnormal patterns reflect the immediate release of pre-existing motor pathways rather than the development of maladaptive circuitry. In the intact nervous system, corticospinal pathways provide selective control over motor output and regulate the expression of more diffuse descending pathways, including reticulospinal projections. Loss of corticospinal input may therefore reduce this regulatory control, allowing less fractionated motor commands to emerge when voluntary movement returns. One hypothesis proposes that abnormal co-activation after corticospinal tract damage is due to reliance on use of residual tracts such as the contralesional corticoreticulospinal tract [17]. However, recent work suggests that components of the obligatory flexor synergy involve spinal circuitry rather than solely cortical mechanisms [18]. Also, epidural stimulation, which targets spinal inhibitory mechanisms, can reduce co-activation between agonists and antagonists and improve reaching performance in chronic stroke survivors [19]. The rapid effect of these interventions could suggest that abnormal co-activation may reflect impaired inhibitory control that remains modifiable.

Previous kinematic studies have suggested that abnormal interjoint coordination may develop early after stroke. In rodents, abnormal shoulder–elbow coordination appeared within 5–14 days after cortical lesioning [10]. In humans, reaching kinematics was characterized by some abnormal elbow and shoulder flexor coupling within the first two months of stroke [9]. Our results extend those kinematic studies by demonstrating that the underlying muscles abnormally co-activate within two weeks of stroke onset [20].

### Implications for rehabilitation

The presence of abnormal co-activation within the first 2 weeks after stroke suggests that rehabilitation begins in a motor system that is already exhibiting impaired coordination. Conventional strengthening and task practice may therefore occur in the context of abnormal muscle coupling, potentially reducing the benefit due to inefficient and abnormal movement patterns. This provides a rationale for combining conventional rehabilitation with interventions that specifically target abnormal co-activation.

Our studies using myoelectric computer interface training [14] have demonstrated that the ability to reduce abnormal co-activation between selected muscle pairs in chronic stroke survivors and improve arm function [7], [12], [13]. This study suggests that such interventions might be more effective if delivered during the subacute phase, and thus warrants further investigation.

### Limitations

This study did have some limitations. First, the study was cross-sectional rather than longitudinal, so we could not track within-subject evolution of abnormal synergies from the subacute to chronic stage. In addition, the subacute and chronic cohorts were recruited using different eligibility criteria, and therefore may not represent exactly the same underlying population. Although most chronic participants with substantial upper-extremity impairment would likely have met the subacute eligibility criteria early after stroke, unmeasured differences between the cohorts cannot be excluded and may have influenced the comparison. The similarities we observed between subacute and chronic groups suggest stability, but future longitudinal work is needed to confirm whether early abnormalities predict long-term patterns. Second, our analysis focused on a somewhat constrained reaching task (although substantially more kinematically complex than many prior studies of synergies, e.g., [9]) while recording from a limited set of nine muscles. Including trunk, hand, and wrist muscles in future EMG recordings could provide a more complete picture of whole-limb modular control. Third, the ipsilesional arm was used as a reference rather than a separate healthy control group. While this is common [8], [15], prior work suggests that ipsilesional control is often close to normal, but subtle bilateral effects of stroke cannot be ruled out [22], [23]. Nevertheless, using each participant’s ipsilesional arm as the reference has the advantage of controlling for individual differences in anatomy and motor strategies [16], [24]. Finally, synergy identification depends on several analytic choices, including the number of modules, normalization, and the similarity metrics used. We used conservative VAF criteria and randomization thresholds similar to those in published synergy work [6], but it remains possible that different computational approaches would reveal additional temporal or spatial components [25], [26].

## Conclusions

In summary, this study demonstrates that abnormal arm muscle co-activation is already present by 2 weeks after stroke and remains abnormal in the chronic stage. A deltoid-dominant synergy with strong anterior–posterior co-activation emerges early, and elbow flexors are recruited abnormally at reach onset. These findings highlight the need for early interventions targeted at abnormal co-activation after stroke.

## Data Availability

All data used in the present study will be made available upon reasonable request to the authors upon publication in a peer-reviewed journal.

## Abbreviations

EMG: Electromyography
FMA UE: Fugl Meyer Assessment – Upper Extremity
WMFT: WMFT Motor Function Test
MAS: Modified Ashworth Scale
SAFE: Shoulder Abduction–Finger Extension score
VAF: Variance Accounted For
IRB: Institutional Review Board
SENIAM: Surface Electromyography for the Non Invasive Assessment of Muscles
AD: Anterior deltoid
MD: Middle deltoid
PD: Posterior deltoid
BI: Biceps brachii
BRD: Brachioradialis
TRIlong / TRI long: Triceps brachii long head
TRIlat / TRI lat: Triceps brachii lateral head
Pec / PECclav: Pectoralis major (clavicular head)
Trap / TRP: Trapezius
ANOVA: Analysis of Variance
Tukey–Kramer: Tukey–Kramer post hoc test

## Acknowledgements

We gratefully acknowledge the participants for their time and commitment to this study. We also thank the members of our research team and the occupational therapists who contributed to data collection and clinical assessments. Special thanks to Veronica Rowe for training our occupational therapists in outcome evaluation procedures.

